# ‘I was an accident’. Long-term effects of unintended pregnancy on children: Findings from the Dutch prospective birth-cohort Amsterdam Born Children and their Development study

**DOI:** 10.1101/2024.01.02.23300277

**Authors:** Wieke Y. Beumer, Marjette H. Koot, Tanja Vrijkotte, Tessa J. Roseboom, Jenneke van Ditzhuijzen

## Abstract

**Background:** Several studies investigated short-term risks of children born from unintended pregnancies, however evidence about long-term risks is missing. This study aimed to examine whether children born from unintended pregnancies experience psychosocial problems up into adolescence.

**Methods:** This study is based on the birth cohort study ‘Amsterdam Born Children and their Development’ (*n* = 7784). Unintended pregnancy was measured as a multidimensional construct. Children’s psychosocial problems were measured with the Strengths and Difficulties Questionnaire, at 5-6, 11-12 and 15-16 years old. Multiple Structural Equation Models were analyzed, examining associations between unintended pregnancy and children’s psychosocial problems, while controlling for co-occurring risks. Mediating effects via maternal mental health and bonding were assessed.

**Results:** Pregnancy mistiming was a significant predictor of internalizing (β = .10, *p* < .001) and externalizing problems (β = .07, *p =* .006) and unwanted pregnancy of internalizing problems (β = .13, *p* < .001) at 5-6 years. These associations were mostly mediated by maternal mental health and poorer maternal bonding. Associations were no longer present at 11-12 and 15-16 years.

**Conclusion:** Children born from unintended pregnancies experience more psychosocial problems at 5-6 years, but no longer in adolescence. Unintended pregnancies often coincide with maternal mental health problems, and the associations between unintended pregnancy and children’s psychosocial problems are influenced by maternal mental health and poorer bonding. Therefore it is important to improve maternal mental health and bonding for the benefit of both mother and child, rather than on the isolated effect of unintended pregnancy per se.

**Key messages:** *What is already known on this topic:* Of the few studies that have been done, most showed that unintended pregnancy is associated with more psychosocial problems of children later in life, but others found no evidence. Especially evidence about long-term effects on children is missing.

*What this study adds:* People who carry a more unintended pregnancy to term experience more mental health issues and poor bonding to their child, which in turn negatively affects their young child. However, when children are older (in adolescence), they no longer face a higher risk of psychosocial problems. This was studied in a context with relative liberal abortion laws and context (i.e., the Netherlands).

*How this study might affect research, practice or policy:* It is important to build on maternal mental health and bonding when wanting to reduce children’s psychosocial problems, instead of focusing on the unintended pregnancy per se.

**Competing Interest Statement:** The authors have declared no competing interest.

## Introduction

Unintended pregnancy is a common phenomenon worldwide (1). Despite being unintended, some result in the birth of much wanted and loved children, some are carried to term despite being unwanted, and some are terminated. It is estimated that around 50% of unintended pregnancies are carried to term (1). There continues to be public concern about the potential harmful consequences of unintended pregnancy on families, and children in particular (2, 3). Thus, it is important to understand the potential impact of unintended pregnancy, to provide meaningful insights for public health and policy makers.

Gipson et al. (3) showed in their review assessing the effects of unintended pregnancy on the health of children, that most research focused on short-term outcomes, specifically mostly on physical health outcomes (like preterm birth). Research into longer-term effects on children’s psychosocial problems is scarce, and findings are inconsistent (3). Of the few results, most showed that unintended pregnancy is associated with more psychosocial problems of children (4, 5), even when they grow into adults (6, 7). However, others found no evidence (8). These mixed findings may result from varying outcomes and definitions of ’unintended pregnancy’ (9, 10), study locations with differing abortion laws, or the focus on populations with relatively low socioeconomic circumstances (3).

Besides inconsistent results, previous research into potential indirect effects explaining impact of unintended pregnancy on children is lacking. People who carried an unintended pregnancy to term, reported more mental health problems later in life (11, 12). Concurrently, maternal mental health has been found to affect children’s psychosocial problems (13). Moreover, people carrying an unintended pregnancy to term reported poorer maternal bonding (14), which might in turn influence the child (15). Thus, an indirect effect of unintended pregnancy on children via maternal mental health and bonding problems is plausible, but this remains unstudied up to now.

The current study aims to fill these research gaps by investigating the association between unintended pregnancy and children’s psychosocial problems over time up into adolescence, and examining the indirect effect via maternal mental health and bonding problems. Associations are investigated while taking the multidimensionality of unintended pregnancy into account (9, 10). Further, we aimed to isolate the effect of being “unintended” by controlling for multiple co-occurring risks for children’s psychosocial problems. This is investigated in a prospective longitudinal birth cohort, assessing possible effects for children growing up in a more abortion liberal context.

## Methods

### Participants and procedure

This study is part of the birth cohort study Amsterdam Born Children and their Development’ (ABCD) (16). The overarching aim of this study is to explore the health and development of children and their families born in Amsterdam, the Netherlands. 12,373 pregnant women were invited to participate during their initial obstetric care visit between 2003-2004. Around 16 weeks gestation, women subsequently received additional written information at home. Those willing to participate signed the informed consent form, completed the questionnaire, and returned it by post. At each subsequent data collection point, informed consent was obtained.

Among the pregnant women recruited, 8,266 participated in the study (response rate 67%), while selection bias was reduced to a minimum (17). Inclusion criteria were being pregnant currently and residence in Amsterdam. In the current study, twins (*n* = 135) and/or children with major congenital diseases (*n* = 162) were excluded. Moreover, only women who were older than 16 years during their pregnancy were included, resulting in a sample size of 7,784 participants. A post hoc statistical power analysis for testing a covariance structure model using Root Mean Square Error of Approximation (RMSEA) was conducted (18), estimating the power for a given RMSEA (Null RMSEA = .05, alternative RMSEA = .1), sample size (*n* = 7,784), and an alpha of .05. Results indicated an adequate sample size (power > .9).

Data were anonymously collected at four measurement phases: (1) antepartum (around 16 weeks gestation), (2) preschool age (5-6 years), (3) early adolescence (11-12 years), (4) middle adolescence (15-16 years) (Figure 1). All measurements consisted of self-reported questionnaires filled out at home. In each questionnaire, it was clearly stated that data were processed confidentially (16).

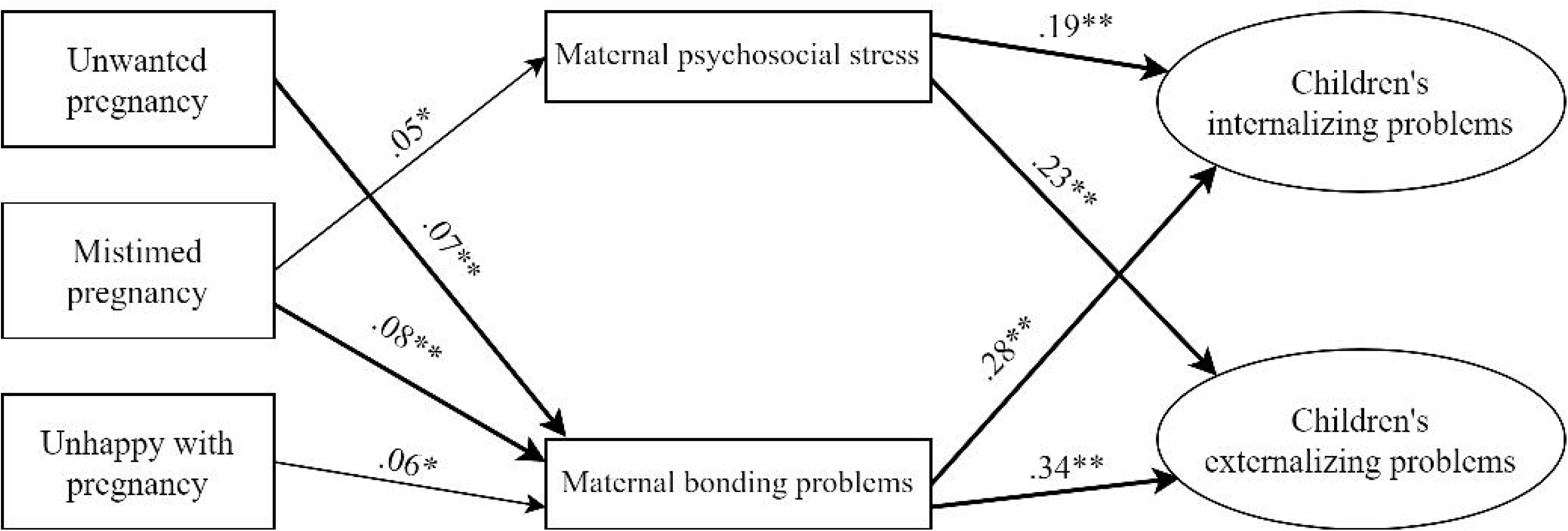

### Outcome measures

Children’s psychosocial problems were measured with maternal-reported subscores of the Strengths and Difficulties Questionnaire (SDQ) (Figure 1) (19). In the present study, the SDQ scores were acceptably reliable (Cronbach’s alpha: .752 (preschool age); .806 (early adolescence); .784 (middle adolescence)). Secondary outcome analyses were performed with child-reported SDQ scores in middle adolescence.

### Predictor variables

#### Pregnancy intentions

During pregnancy, pregnancy intentions were measured as a multidimensional construct, based on maternal-reported data on the extent of pregnancy mistiming, unwantedness and unhappiness. They were measured with an item each: ‘*This pregnancy happened too soon*’ (mistiming), ‘*I did not want to be pregnant (anymore*)’ (unwantedness), and *‘I am happy to be pregnant’* (unhappiness; recoded). Items were rated on a 4-point Likert scale ranging from 0 (*definitely not true*) to 3 (*very true*).

#### Mediators: Maternal factors

Mediators were measured in each corresponding measurement phase (preschool, early- and middle adolescence). First, maternal bonding problems were measured with the bonding subscale of the Nijmeegse Ouderlijke Stress index (NOSI-K) (20), consisting of 9 items rated on a 4-point Likert scale ranging from 0 (*definitely not true*) to 3 (*very true*). A total score was calculated, with higher scores indicating more bonding problems. The NOSI-K showed good reliability (Cronbach’s alpha = .850).

Second, maternal psychosocial distress was measured with the reliable and validated Dutch version of the Depression Anxiety Distress Scale (DASS-21) (21, 22). The DASS-21 measured symptoms of depression, anxiety and distress over the preceding week on a 4-point Likert scale ranging from 0 (*never or rarely*) to 3 (*very often*). A total score was calculated, with higher scores indicating more psychosocial distress. The DASS-21 showed excellent reliability (Cronbach’s alpha: .900).

#### Controls

First, confounding antepartum maternal variables should be considered when studying risks of unintended pregnancy (3). For instance, younger maternal age and lower socioeconomic status has been associated to both unintended pregnancy and children’s psychosocial problems (8). Thus, we controlled for maternal age and years of education.

Further, maternal antepartum mental health and substance use, are associated to both unintended pregnancy (2) and children’s psychosocial problems (6). Hence we controlled for maternal antepartum mental health in the current study. Depressive symptoms were measured with the Center for Epidemiological Studies Depression scale (CES-D) (23). Anxiety was assessed with the State-Trait Anxiety Inventory State form (STAI-S) (24). For both, a total score was calculated, with higher scores indicating more symptoms. Moreover, a total ‘antepartum risk behaviour’ variable was calculated, based on maternal smoking, drinking and drug use in pregnancy. Total scores ranged from 0 to 3, with higher scores indicating more risk behaviour.

Moreover, co-occurring child risk factors may also independently affect children, such as the child’s birth sex, living situation (e.g., having divorced parents), and experiences with adverse life events (ACE) (such as the death of a family member) (25–27). Thus, we controlled for birth sex and at every measurement for the age and living situation of the child (with both parents (together, or co-parenting) vs. not with both parents). Further, since the number of experienced ACE’s is associated with children’s psychosocial problems (25), a total ‘score’ was computed.

### Statistical analyses

All statistical analyses were performed in R studio (28). Associations between unintended pregnancy and children’s psychosocial problems were investigated using Structural Equation Modelling (SEM). Models were considered adequately fitted when the RMSEA was smaller than .05, and the Comparative Fit Index (CFI) was larger than .90 (29). To account for non-normality in the data, Robust Maximum Likelihood (MLR) was used, by computing robust standard errors (30). Missing data were handled using Full Information Maximum Likelihood (FIML) estimation, providing more reliable results compared to other ways of handling missing data (31).

Analyses were performed per measurement (preschool age, early- and middle adolescence). First, a confirmatory factor analysis (CFA) of the latent factor structure of the outcome variables was conducted. After confirming adequate fit, a path model was estimated, indicating associations between unintended pregnancy dimensions and children’s internalizing- and externalizing behaviour problems. We controlled for co-occurring factors by adding them as predictor variables (Figure 1). Regression coefficients were assumed to be significant when they had a *p*-value below .05. We additionally examined the associations between unintended pregnancy and children’s psychosocial problems, as reported by the child in adolescence. Lastly, using SEM as well using the step-by-step approach (32), we tested the mediating effect of maternal psychosocial distress and bonding problems.

### Non-response analyses

Participants who were lost to follow up were compared to those who responded to each measurement. Detailed results are reported in an earlier published manuscript (11). In short, compared to responders, dropouts reported more antepartum mental health problems and more unintended pregnancies.

## Results

### Descriptive analyses

Data of 7,784 participants and their children were included (Figure 1). Participants were between 16-46 years old during pregnancy. Other participant characteristics are described in Table 1. Table 2 shows correlations between the variables of interest. Most importantly, people who reported more pregnancy unintendedness, also reported more internalizing problems of their children at 5-6, 11-12 and 15-16 years. Further, results showed that all three aspects of unintended pregnancy were positively correlated to children’s externalizing problems at 5-6 years. In contrast, only pregnancy unhappiness and mistiming were positively correlated to children’s externalizing problems at 11-12 years. And in late adolescence (15-16 years), unintended pregnancy was no longer correlated to children’s externalizing problems.

**Table 1.** Participant characteristics (*n* =7,784).

| **Antepartum maternal characteristics** |  |  |
| --- | --- | --- |
| Age ($M$, $SD$) | Years | 30.73 (5.22) |
| Risk behaviour (yes: $n$, %) | Smoking | 454 (5.8) |
|  | Drinking alcohol | 1502 (19.3) |
|  | Drug use | 69 (0.9) |
| State anxiety symptoms ($M$, $SD$) | STAI-score | 38.35 (10.25) |
| Depressive symptoms ($M$, $SD$) | CES-D score | 12.67 (8.6) |
| Pregnancy unhappiness ($n$, %)<br>*I am happy to be pregnant* | *Completely true* | 6279 (80.6) |
|  | *True* | 1389 (17.8) |
|  | *Not true* | 70 (0.9) |
|  | *Completely not true* | 14 (0.2) |
|  | Missing | 32 (0.4) |
| Pregnancy unwantedness<sup>b</sup> ($n$, %)<br>*I did not want to become pregnant (anymore)* | *Completely true* | 162 (2.1) |
|  | *True* | 419 (5.4) |
|  | *Not true* | 1548 (19.9) |
|  | *Completely not true* | 5623 (72.2) |
|  | Missing | 32 (0.4) |
| Pregnancy mistiming<sup>b</sup> ($n$, %)<br>*This pregnancy came too soon* | *Completely true* | 335 (4.3) |
|  | *True* | 885 (11.4) |
|  | *Not true* | 2904 (37.3) |
|  | *Completely not true* | 3350 (45.1) |
|  | Missing | 110 (1.4) |
| **Children's characteristics** |  |  |
| Sex child ($n$, %) | Boy | 3908 (50.2) |
|  | Girl | 3846 (49.4) |
|  | Missing | 30 (0.4) |
| ACE at 15-16 years ($n$, %)<sup>c</sup> | None | 83 (4.8) |
|  | 1 | 225 (13.1) |
|  | 2 | 313 (18.2) |
|  | 3 | 369 (21.4) |
|  | 4 | 330 (19.2) |
|  | >4 | 403 (23.4) |
| Living situation at 15-16 years ($n$, %) | With both parents (incl. co-parenting) | 1819 (86.9) |
|  | Not with both parents | 275 (13.1) |

M, mean; SD, standard deviation; STAI, State-Trait Anxiety Inventory, CES-D;
Center for Epidemiologic Studies Depression Scale; ACE; Adverse Childhood Experiences
<sup>b</sup> Pregnancy intention dimensions were measured on a 4-point Likert scale (0-3), with a higher score indicating more unintendedness<sup>c</sup> In the SEM analyses, ACE was measured as a total sum score, with a higher score indicating more experiences with ACE

**Table 2.**
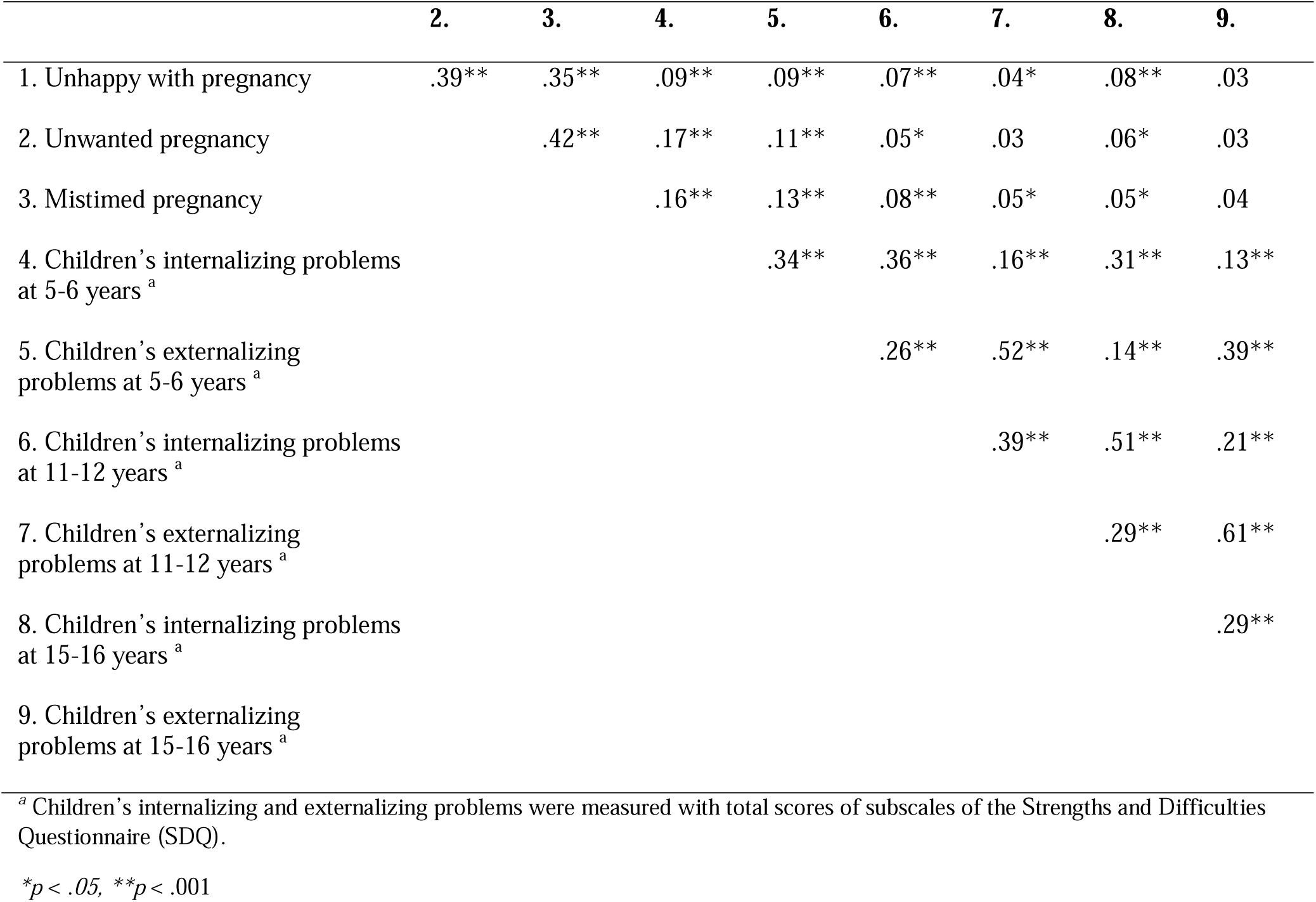
Bivariate correlations between main variables of interest.

|  | 2. | 3. | 4. | 5. | 6. | 7. | 8. | 9. |
| --- | --- | --- | --- | --- | --- | --- | --- | --- |
| 1. Unhappy with pregnancy | .39** | .35** | .09** | .09** | .07** | .04* | .08** | .03 |
| 2. Unwanted pregnancy |  | .42** | .17** | .11** | .05* | .03 | .06* | .03 |
| 3. Mistimed pregnancy |  |  | .16** | .13** | .08** | .05* | .05* | .04 |
| 4. Children's internalizing problems at 5-6 years <sup>a</sup> |  |  |  | .34** | .36** | .16** | .31** | .13** |
| 5. Children's externalizing problems at 5-6 years <sup>a</sup> |  |  |  |  | .26** | .52** | .14** | .39** |
| 6. Children's internalizing problems at 11-12 years <sup>a</sup> |  |  |  |  |  | .39** | .51** | .21** |
| 7. Children's externalizing problems at 11-12 years <sup>a</sup> |  |  |  |  |  |  | .29** | .61** |
| 8. Children's internalizing problems at 15-16 years <sup>a</sup> |  |  |  |  |  |  |  | .29** |
| 9. Children's externalizing problems at 15-16 years <sup>a</sup> |  |  |  |  |  |  |  |  |

<sup>a</sup> Children's internalizing and externalizing problems were measured with total scores of subscales of the Strengths and Difficulties Questionnaire (SDQ).
\* $p < .05$ , \*\* $p < .001$

### Direct over-time associations between unintended pregnancy and children

The over-time associations between unintended pregnancy dimensions and children’s internalizing- and externalizing problems were analysed per measurement (preschool age, early- and middle adolescence). To establish adequate model fit per measurement, the modification indices suggested covariance between different items within subscales. These covariances theoretically made sense, since those items were derived from the same construct. Thus, models were adjusted accordingly, resulting in improved, adequate model fit in each model (preschool age: CFI = .920, RMSEA = .017, early adolescence: CFI = .912, RMSEA = .017, middle adolescence: CFI = .904, RMSEA = .015) (29).

Pregnancy mistiming was a significant predictor of more internalizing and externalizing behavior problems of children at preschool age (β = .10, *p* < .001; β = .07, *p =* .006, respectively). Effect sizes are small (33). Further, unwanted pregnancy was positively associated to internalizing problems (β = .13, *p* < .001), though not to externalizing problems. Moreover, pregnancy unhappiness did not significantly predict internalizing and externalizing behavior problems (Table 3).

**Table 3.**
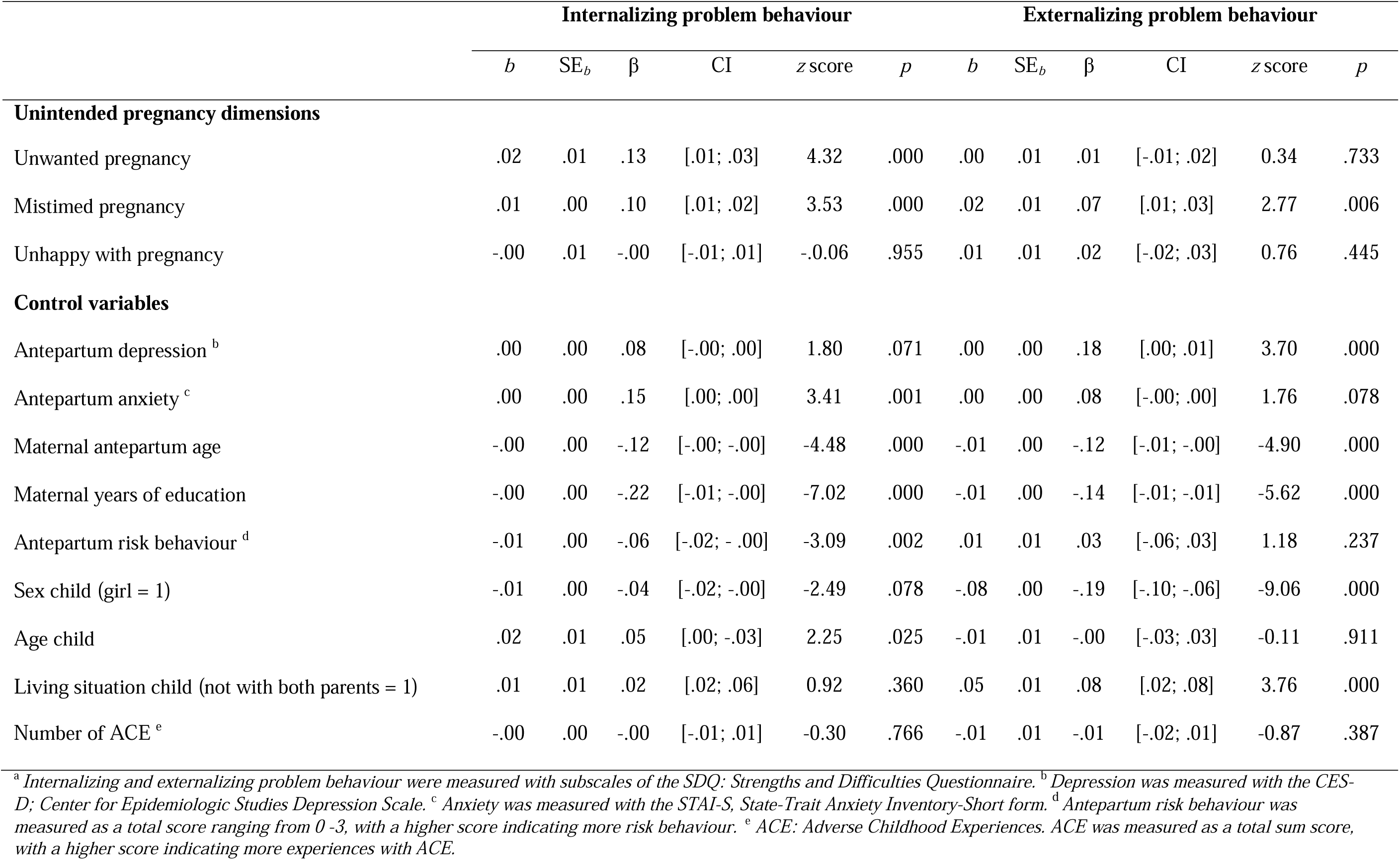
Structural Equation Modelling regression results of the association between predictor variables and children’s internalizing- and externalizing problems in preschool age (5-6 years old)

In contrast, both in early and middle adolescence, not one dimension of unintended pregnancy was significantly associated to internalizing- and externalizing problems while controlling for co-occurring risks. Additional sensitivity analyses with child-reported SDQ scores as a secondary outcome in middle adolescence, showed the same absence of associations between unintended pregnancy dimensions and children’s internalizing- and externalizing problems.

### Indirect associations via maternal mental health and bonding problems

Since the current study did not reveal a direct effect of unintended pregnancy dimensions on children in adolescence, only mediation results for children at preschool age were studied. This mediation model had an adequate model fit (CFI = .908, RMSEA = .019). All effects were tested simultaneously, to test if the mediation effect is independent of the effects of the other mediator and controls.

Results showed a significant mediating effect of maternal bonding on the associations between unintended pregnancy aspects and children’s internalizing and externalizing problems (Figure 2). Unintended pregnancy dimensions were positively associated to maternal bonding problems (unhappy: β = .06, *p* = .008; unplanned: β = .08, *p* < .001; unwanted: β = .07, *p* = .001). Effect sizes are small (33). In turn, maternal bonding problems positively affected children’s internalizing- and externalizing behavior problems (β = .29, *p* < .001; β = .35, *p* < .001, respectively). Effect sizes are medium (33).

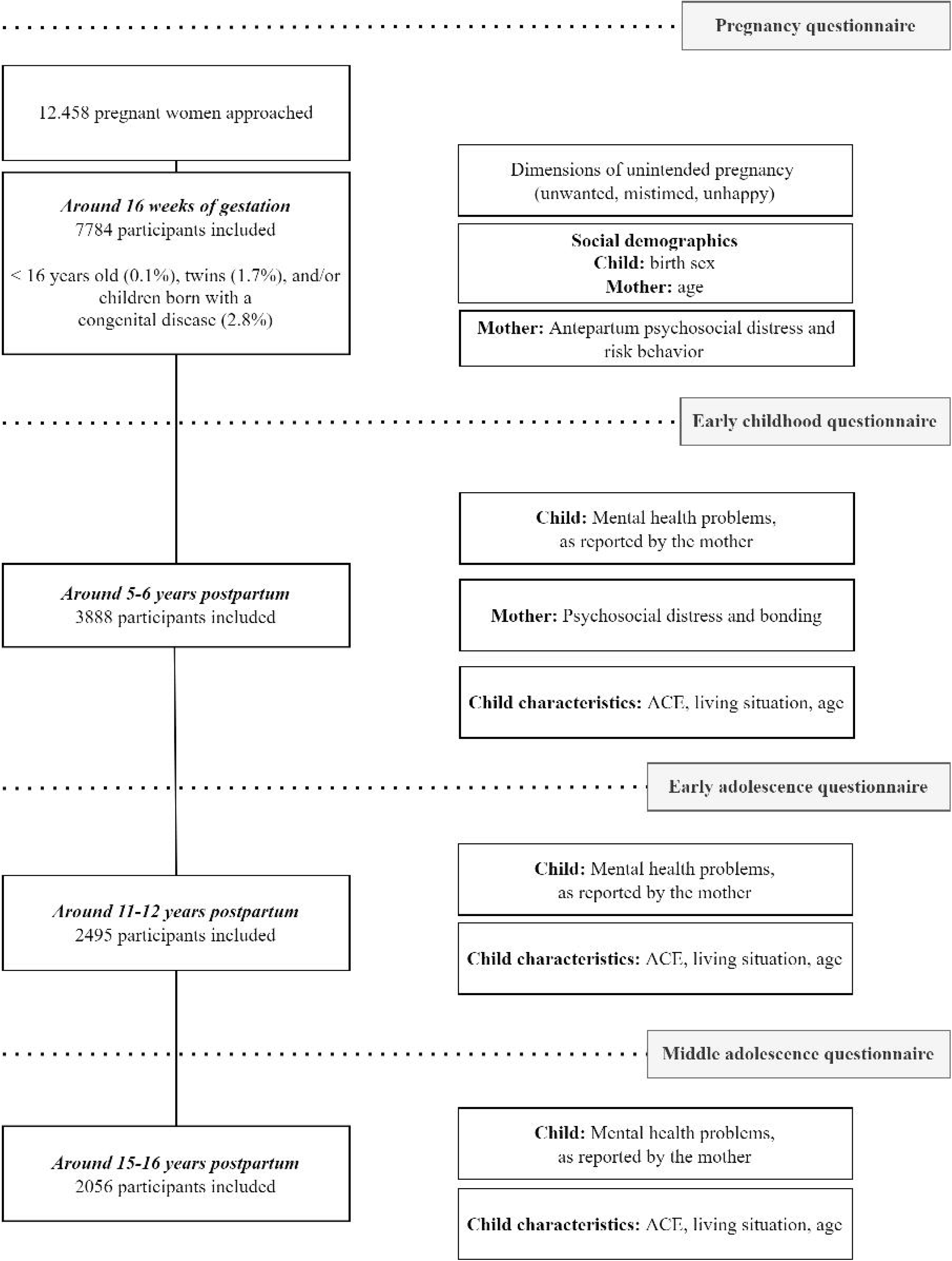

Results are more complex regarding the mediating role of maternal mental health, since maternal mental health was only a significant mediator on the association between pregnancy mistiming and children’s internalizing and externalizing problems (Figure 2). Mistimed pregnancy positively affected maternal psychosocial distress (β = .05, *p* = .016) and in turn, this positively affected children’s internalizing- and externalizing behavior problems (β = .20, p < .001; β = .23, p < .001 respectively). Effect sizes are small (33).

Notably, even after accounting for the mediators and controls in the model, some direct associations between unintended aspects and children’s internalizing and externalizing problems were still present. Pregnancy mistiming and unwantedness were still significant predictors of children’s internalizing problem behavior (β = .07, *p* = .016; β = .15, *p* <. 001 respectively), indicating small effect sizes (33) and complementary mediation (32).

## Discussion

Many children are born from unintended pregnancies (1), and are sometimes labeled as ‘accidents’. The current study investigated whether these children experienced more psychosocial problems (up into adolescence), and whether this was influenced by their mother’s mental health and bonding. We only found evidence for a small increase in psychosocial problems in preschool age (as reported by their mothers), but no longer in adolescence.

The current study results show that dimensions of unintended pregnancy are associated with more psychosocial problems of children in preschool age, reported by their mothers, which is consistent with results of previous studies (4, 5). Most of this association was explained by concurrent mental health and bonding problems of the mother. People who carried an unintended pregnancy to term reported poorer bonding and mental health, which in turn was associated to psychosocial problems of the child . This is in line with previous studies, which found that people carrying an unintended pregnancy to term reported poorer maternal bonding (14) and more mental health problems in the first years postpartum (11). Thus, current results stress the importance of maternal mental health for children, which appeared to be a stronger factor contributing to children’s psychosocial problems than pregnancy unintendedness. Hence, it might be fruitful to invest in extra support promoting the mental health and bonding of people carrying an unintended pregnancy to term, which in turn might also positively affect their children.

The increase in psychosocial problems among children born from unintended pregnancies could not be detected anymore in adolescence. The first peaking, but then disappearing effect of unintended pregnancy on children is in line with trends found in other studies examining over-time effects of unintended pregnancy (34). Further, this is in line with previous results showing disappearing risks of unintended pregnancy on *maternal* mental health over time (11).

However, in contrast to current results, other studies indicated a higher risk of problems for children born from an unintended pregnancy up into adolescence and adulthood (3, 6, 7). This inconsistency in findings can be explained by the fact that previous studies were mostly done in populations in which women were not legally allowed to have an abortion, or were denied one (5, 7). In the current study’s context, people who carried a strongly unwanted pregnancy may have chosen to terminate this pregnancy, and therefore were not part of the current study sample. People who had an abortion, indeed reported more pregnancy unintendedness compared to people who chose to carry an unintended pregnancy to term in the Netherlands in a previous study (35). We do not know what the results would have been if these highly unwanted pregnancies were carried to term, and hence part of this study.

Another explanation for inconsistent findings in comparison to previous studies, is that different definitions of unintended pregnancy are used. Pregnancy intentions are often operationalized in a dichotomous way (being either intended *or* unintended). However, pregnancy intentions are a complex construct with a lot of factors involved (9, 10). The current study measured pregnancy intentions as a multidimensional construct, thereby providing more insightful results. Results showed that pregnancy mistiming was the strongest predictor of children’s psychosocial problems. This might also be explained by the retrospective measurement of the pregnancy intention dimensions in the current study. As the pregnancy continutes, people tend to perceive their pregnancy to be more wanted and state to be more happy with their pregnancy (35, 36).

Moreover, inconsistent findings can be explained by the fact that in contrast to most previous studies (8), the current study controlled for the effects of co-occurring risk factors. Current results showed the influence of personal and family background variables on children’s psychosocial problems, compared to the effects of the unintended pregnancy itself. This indicates the relevance of considering co-occurring risk factors when investigating (consequences of) unintended pregnancy (37).

We acknowledge several limitations of this study. First, we cannot eliminate the risk that selection bias in participants who dropped out our cohort data affected our results. Cohort studies are prone to selective dropout and hence are likely to underestimate prevalence of unintended pregnancy and mental health problems (38). A second limitation might be that the children’s psychosocial problems was reported by the mother in the current study. However, mothers with more mental health problems perceive their children as having more psychosocial problems (39). Nevertheless, current study’s additional analyses investigating adolescent-reported psychosocial problems, showed the same pattern of results. Lastly, since previous studies showed the important influence of paternal mental health on children (40), future studies are advised to incorporate the role of the co-parent.

Despite these limitations, this study was the first to investigate multiple long-term associations between unintended pregnancy and children’s psychosocial problems. Our findings were derived from a large-scale birth cohort. Hence, results might be representative for the Dutch population, and other countries with similar abortion policy and cultures. Further, in line with earlier recommendations (3, 9), the design of the current study made it possible to consider the complexity of pregnancy intentions, by taking into account different dimensions of unintended pregnancy and consequently providing more meaningful results. Moreover, associations were investigated while taking into account other important co-occurring risks.

## Conclusions

People who carry a more unintended pregnancy to term experience more mental health issues and poor bonding to their child, which in turn negatively affects their young child. This implies the importance of focusing on maternal mental health and bonding when wanting to reduce children’s mental psychosocial problems, instead of focusing on the unintended pregnancy per se.

## Data Availability

All data produced in the present study are available upon reasonable request to Tanja Vrijkotte

